# Helium optically pumped magnetometers can detect epileptic abnormalities as well as SQUIDs as shown by intracerebral recordings

**DOI:** 10.1101/2023.10.24.23297371

**Authors:** Jean-Michel Badier, Denis Schwartz, Christian-George Bénar, Khoubeib Kanzari, Sébastien Daligault, Rudy Romain, Sergey Mitryukovskiy, William Fourcault, Vincent Josselin, Matthieu Le Prado, Julien Jung, Augustin Palacios-Laloy, Romain Carron, Fabrice Bartolomei, Etienne Labyt, Francesca Bonini

## Abstract

SQUID-based magnetoencephalography has been shown to improve the diagnosis and surgical treatment decision for presurgical evaluation of drug-resistant epilepsy. Still, its use remains limited due to several constraints such as cost, fixed helmet size and obligation of immobility. A new generation of sensors, the optically pumped magnetometers (OPMs), could overcome these limitations. In this study, we validate the ability of innovative Helium-based OPM (^4^He-OPM) sensors to record epileptic brain activity thanks to simultaneous recordings with intracerebral EEG (stereotactic EEG, SEEG). We recorded simultaneous SQUIDs-SEEG and 4He-OPM-SEEG signals in one patient during two sessions. We show that epileptic activities on intracerebral EEG can be recorded by OPMs with a better signal-to noise ratio than classical SQUIDs. The OPM sensors open new venues for the widespread application of magnetoencephalography in the management of epilepsy and other neurological diseases and fundamental neuroscience.

**Significance Statement:** We performed a simultaneous recording of Helium-based Optically Pumped Magnetometers (OPM) and intracerebral EEG and validate for the first time OPM results with signals recorded directly within the brain. We demonstrate that epileptic abnormalities seen on intracerebral electrodes are detected by OPMs with a better signal-to noise ratio than classical magnetoencephalography. This represents a significant step towards the validation of OPM-based recordings for epilepsy diagnosis and for clinical and neuroscience research.

## Introduction

Magnetoencephalography (MEG) is a non-invasive, millisecond-resolution source imaging technique for recording and localizing brain signals. The analysis of magnetic brain activity allows for a deeper understanding of the neural substrates of brain pathologies. In the management of drug-resistant epilepsy, MEG contributes to the success of surgical treatment through non-invasive localization of interictal epileptic discharges (IEDs) and can also guide the implantation of depth electrodes when invasive recordings are required [Fischer et al., 2005; Murakami et al., 2016].

Still, the spread of MEG is strongly limited by the constraints imposed by the use of superconducting quantum interference devices (SQUIDs)[Cohen, 1972; Hämäläinen et al., 1993], requiring cooling at very low temperature (4.2 K). Thus, the sensors are isolated and enclosed in a fixed array inside a rigid dewar, at least 3 cm from the brain. The critical consequences are a substantial reduction in the magnetic signal amplitude, inhomogeneous coverage, and the need for total immobility during signal acquisition.

The emergence of a new generation of MEG sensors, the optically-pumped magnetometers (OPM)[Budker and Romalis, 2007], could overcome these limitations. OPM are quantum sensors that exploit the interaction between an atomic gas and laser light to obtain very precise measurements of tiny magnetic fields.. OPM do not require extreme cooling, and can be placed near the scalp in a wearable system thus allowing subject’s movement [Boto et al., 2017; Iivanainen et al., 2017; Johnson et al., 2013] [Boto et al., 2018; Boto et al., 2022; Seymour et al., 2021]. The development and improvement of this new technology has taken a quantum leap in recent years, with evolution in miniaturization and sensitivity (for complete reviews see [Brookes et al., 2022; Labyt et al., 2022; Tierney et al., 2019]). The first commercially available OPMs, based on Alkali atoms[Budker and Romalis, 2007], have a sensitivity of 20fT/rtHz in tri-axis mode compared to 5 fT/rtHz for SQUID sensors. However, their placement close to the scalp allows a 3-8 fold increase in signal power [Budker and Romalis, 2007; Iivanainen et al., 2017; Johnson et al., 2013], with a significant neuromagnetic signal enhancement compared to SQUIDs, namely for superficial sources[Boto et al., 2016; Boto et al., 2017; Iivanainen et al., 2017].

More recently, new OPMs based on Helium atoms (^4^He-OPMs) have been developed[Beato et al., 2018], having a large dynamic range (up to 250 nT) and a large frequency bandwidth (up to 2kHz) with negligible heat dissipation (10 mW per sensor). Their properties allow recording in a standard magnetic shielding room, facilitating data acquisition.

A few studies have shown that alkali OPMs can record IEDs comparable to those observed with SQUID-MEG [Feys et al., 2022] or previously obtained EEG [Vivekananda et al., 2020]. However, to compare OPM to SQUID-MEG, it is necessary to ensure that both modalities are recording equivalent IEDs. By using stereotactic intracerebral-EEG (SEEG) as a groundtruth, simultaneously recorded with OPMs and SQUIDs[Badier et al., 2017], we aimed to demonstrate that ^4^He-OPMs can perform as well as SQUID-MEG but at an expected lower cost and with greater convenience. These unique simultaneous recordings allowed selecting similar IEDs to compare SQUID-MEG and ^4^He-OPM and thus evaluate the Signal to Noise Ratio (SNR) of both techniques.

## Methods

The simultaneous recordings were performed inside a 2-layer μ-metal magnetic shielded-room (MSR) on a patient with drug-resistant focal epilepsy undergoing SEEG. Fourteen intracerebral electrodes with a total of 119 recording contacts were implanted mainly in the left temporal structures (Fig. 1d). These electrodes, and particularly those exploring the anterior medial and lateral temporal regions, disclosed abundant interictal epileptic abnormalities. A first SQUID-MEG/SEEG recording session (Fig. 1a) of 20 minutes at rest was followed by a comparable ^4^He-OPM/SEEG session. The four ^4^He-OPM sensors (2 x 2 x 5 cm) fixed on a rigid helmet (Fig. 1b) were placed over the left central and temporal regions, in contact with the bandage covering the scalp (Fig. 1c, red dots). The four SQUID sensors closest to the four ^4^He-OPMs sensors were selected for analysis and for comparison with the magnetic activity recorded by the OPM sensors (Fig. 1c, green dots). These four SQUIDs were between 2.72 cm and 3.23 cm away from the OPM sensors.

**Fig. 1.**
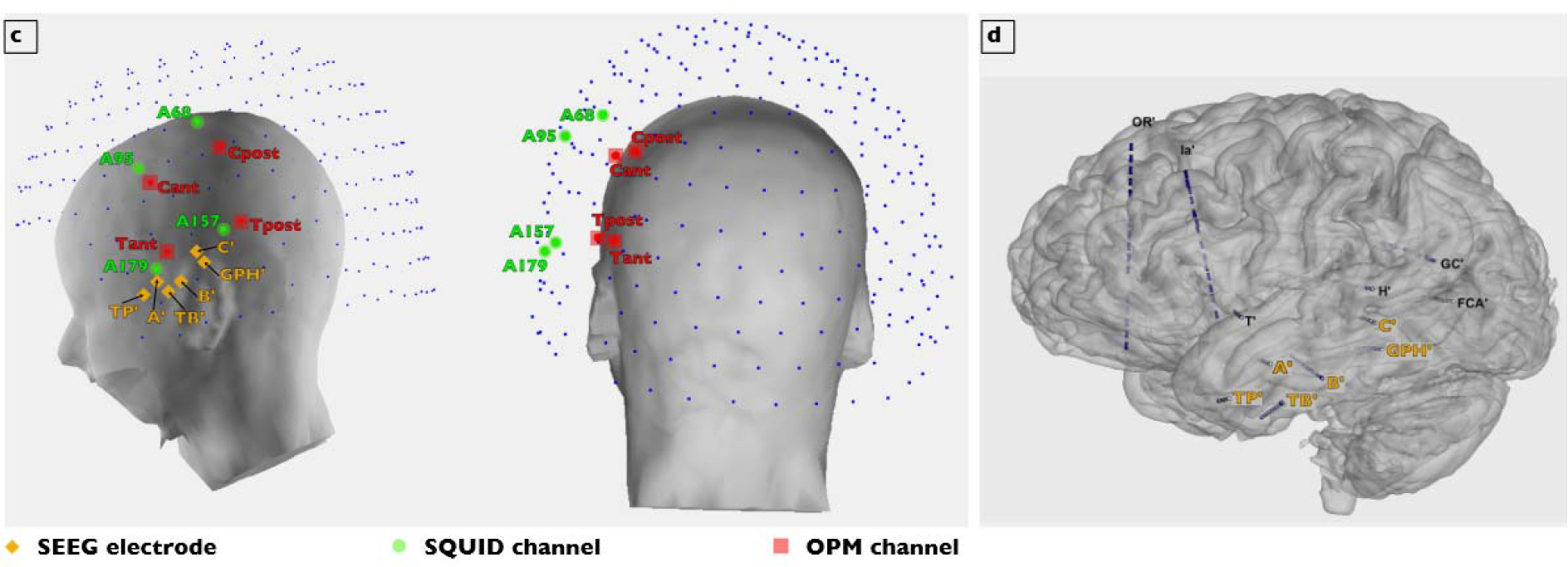
SQUID-MEG, ^4^He-OPM-MEG and SEEG recording setup. a) and b) items have been removed from this MedXriV submission as required for preventing any possible person identification. readers Please contact the corresponding author to request access to these materials. a) Simultaneous SQUID-MEG/SEEG: the classic cryogenic MEG system, measuring 120 cm x 100 cm and weighting about 300 Kg, with 265 SQUID sensors in a fixed array requiring subject immobility during data acquisition. b) Simultaneous ^4^He-OPM-MEG/SEEG: recording configuration composed of 4 sensors integrated in a wearable helmet placed on the scalp and in contact with the bandage covering the SEEG electrodes inserts (the cables and connectors coming out of the helmet are visible). In the top left insert is a picture of the He-OPM sensor. c) 3D reconstruction of the patient’s head from MRI, with He-OPM (in red), SQUID sensors (in blue) and SEEG electrodes entry points (in orange). The 4 SQUID sensors, closest to the OPMs, are coloured in green. Note the distance between the SQUIDs and the scalp (at least 3 cm), while the He-OPM are in contact with it. d) SEEG implantation: 14 intracerebral electrodes, 2 in the right hemisphere (not shown) and 12 in the left hemisphere exploring the whole left temporal structures: A’: amygdala, TP’: temporal pole; TB’: rhinal cortex; B’ anterior hippocampus; C’: posterior hippocampus; GPH’: para-hippocampal gyrus; T’: anterior insula/lateral T1; H’: thalamus/Heschl gyrus gyrus; Ia’: anterior insula/F2; FCA’: lingual gyrus; GC’: posterior cingulate/T1; OR’ orbito-frontal cortex/middle frontal sulcus

### Patient

The patient (age range 31-35 years old) suffered from intractable temporal epilepsy. A stereo-EEG (SEEG) was performed to define the EZ to be removed. Most electrodes (12/14) were aimed at an extensive exploration of the left temporal structures, as well as the anterior insula and the orbito-frontal cortex; two electrodes were implanted in the right anterior temporal region. At the end of the SEEG, the epileptogenic zone network could be defined as involving the left mesial temporal structures including the amygdala, hippocampus, rhinal cortex, left parahippocampal cortex and collateral sulcus (the latter being posterior to the cavernoma) and the left temporal pole. The simultaneous recording session was carried out at the end of the long-term video-SEEG recording, at J11 from electrodes implantation once all clinical data were acquired. The ethical approval was obtained at the XXXX under ID RCB 2020-A01830-39 and the patient gave informed consent.

### SEEG recordings

SEEG exploration was performed using intracerebral multiple contacts electrodes placed intracranially with robotic assistance (ROSA, Zimmer Biomet) and intraoperative Mobius-Airo CT-scan (Stryker, US) verification. Small insertion screws (2023-VG-C-10 or -15, Alcis Besancon) were used to minimize the bulk of the electrodes around the skull and to allow for simultaneous recordings. The electrodes had a diameter of 0.8 mm, contained 10 to 15 contacts. Each contact consists of 2 mm long platinum-iridium and separated from each other by 1.5 mm of insulated material (XXXXX). To accurately define the anatomical position of each SEEG contact along the electrode trajectory, a CT-scan/MRI data fusion was performed using the in-house software XXXX. This Matlab-based tool is able to co-register the MRI to the CT-scan, and to automatically segment and localize depth electrodes contacts by image processing. The signals were formatted in a bipolar configuration keeping only non-contiguous bipolar channels. Recording was made with a BrainProducts BrainAmp DC amplifier. SEEG data have been sampled at 2500Hz.

### SQUID-MEG recordings

The first recording was done with a SQUID MEG system simultaneously with the SEEG (see above) in supine position. MEG Signals were acquired on a 4D Neuroimaging™ 3600 whole head system at a sampling rate of 2034.51 Hz with a total of 248 magnetometers. Additionally, 3 magnetometers and 9 gradiometers were used for noise compensation. The electrocardiographic and the electrooculographic activity were recorded on bipolar EEG channels. SQUID-MEG/SEEG recording was done in supine position.

### ^4^He-OPM recordings

The second recording were done with a prototype of five ^4^He-OPM sensors simultaneously with SEEG, also in supine position. The^4^He-OPMs are sensors that measure the brain magnetic field along the three axes with a continuous self-compensation of the magnetic field on all axes. The magnetic field measurement relies on a measure of the variation of the light absorption caused by the deviation of the electronic spin of ^4^He atoms from the state originally set by a laser pumping. Technical information and physical principles used in our sensors can be found in a previous publication [XXXXX] and are summarized below. The OPM used in this study is based on parametric resonance of helium-4 metastable atoms at near zero magnetic field[Beato et al., 2018; Dupont-Roc, 1971]. The cell containing the ^4^He gas is a cylinder of 1 cm diameter and 1 cm height. This cell, placed at the bottom of the sensor, is surrounded by small 3-axis Helmholtz coils which are used to apply both the RF fields and the compensation fields (see below). A High-Frequency (HF) discharge (excited between 10 and 20 MHz and consuming around 10 mW power) excites the ^4^He atoms from their ground state to the metastable triplet state which has three Zeeman sublevels. A selective optical pumping (with a linearly polarized beam tuned on the D0 line at 1083 nm) is performed to prepare a macroscopic magnetic moment on the gas, which evolves in the magnetic field created by the brain. In our OPMs, to derive a vector measurement of the three components of the magnetic field, two RF fields are applied to the gas: *B*_*Ω*_*cosΩt* along one tangential X axis and *B*_*⍰*_*cos⍰t along* the radial Y axes. Both are orthogonal to polarization of the pump laser beam. Thanks to this scheme first introduced by Dupont-Roc[Dupont-Roc, 1971], three resonance signals are detected on the transmitted pump light at *Ω,⍰*, and *⍰±Ω*. To first order, the amplitude of each resonance is respectively proportional to one of the three components of the magnetic field to be measured (respectively Bx, By, and Bz).

Each sensor is operated in a closed-loop mode on the three axes. This consists in continuously cancelling the three components of the magnetic field by applying an opposite field with the 3-axis Helmholtz coils. The value of each magnetic field component is deduced from the current injected in the compensation coil. In this way the sensor becomes self-calibrated, i.e. its output can only be affected by variations of the transfer function between current and magnetic field set by the coil geometry, and not by other operating parameters (light intensity, HF power…). This closed-loop mode suppresses the cross-axis effects[Cohen-Tannoudji et al., 1970a; Cohen-Tannoudji et al., 1970b; Dupont-Roc, 1970; Dupont-Roc, 1971] by which the measurement of one axis becomes dependent of the field along another axis. This phenomenon has been recently referred as the Cross-Axis Projection Error (CAPE)[Borna et al., 2022], yielding both phase errors and a tilt of the sensing axis. Previous studies with alkali OPM operated in an open-loop mode have characterized an axis tilt of3.3°/nT at low frequencies[Borna et al., 2022] and offsets as small as 1.5 nT resulted in gain errors of ∼ 5%[Boto et al., 2018]. Thereby, ^4^He OPM is the first sensor, to our knowledge, to provide a measurement of the magnetic field components in a closed-loop mode along the three axes, guaranteeing the reliability of the measurement and avoiding any CAPE. Another important advantage of closed-loop operation is the possibility of broadening the dynamic range well above the atomic linewidth. A dynamic range of +/- 250nT is currently achieved for our ^4^He OPMs. The sensitivity of our magnetometer operating in the closed-loop tri-axial mode is better than 50 fT/Hz^1/2^on two of the three axes (the radial and one tangential) with a bandwidth going from DC to 2 kHz.

However, if the closed-loop mode avoids CAPE, it has some unwanted consequences due to the cross-talks that unavoidably exist between the sensors within the OPM array. This problem can be solved by appropriate post-processing as far as the cross-talks are appropriately characterized (for a detailed description, see XXXX). The measured cross-talk matrix for an array of four ^4^He OPM sensors with only 2-mm spacing, which corresponds to an extremely unfavourable situation as compared to a real OPM MEG set-up, revealed low cross-talk (<10%) and showed a good agreement with the estimated matrix from the Biot-Savart calculations. Knowing this cross-talk matrix, minor cross-talk related errors are corrected in the measurement by adequate post-processing. OPM data have been sampled at 11161 Hz and downsampled to 4 kHz.

### OPM, SQUID and SEEG spatial Co-registration

The spatial co-registration was performed for both sessions using a 3D Polhemus digitizer, based on three fiducial markers (nasion, left and right pre-auricular points). The quality of the co-registration was checked using the digitization of the facial mask. Following the standard procedure for 4D Neuroimaging, the position of nasion and both tragi were digitized to allow the construction of the patient frame. The positions of 5 coils located on the subject’s head were also digitized. Activations of these 5 coils before and after recording allowed to determine the location of the MEG sensors within the patient frame. For the ^4^He OPM-MEG/SEEG session, the same three fiducial markers were digitized in the same reference frame. In this case, the sensors were linked to the subject head by the ^4^He OPM headset. This headset was positioned on the subject head prior to the insertion of the ^4^He OPM sensors and the corresponding slots were digitized giving the location of each ^4^He OPM sensor within the same patient reference frame. Finally, both types of sensors were localized in the same reference frame, i.e. the subject head. It was then possible to select the SQUID sensors closest to each ^4^He OPM based on Euclidean distance.

### Data processing

Noise correction: A noise compensation was performed for SQUIDs thanks to recordings from reference sensors (magnetometers and gradiometers). Noise compensation is obtained by subtracting the contribution of the noise measured by the references for each sensor. As there are not enough sensors to perform it, there is no such compensation for the OPM. In that case noise cancellation is only performed by filtering (band-pass filter 2Hz-70Hz and a notch filter at 50Hz). Downsampling and temporal registration: since the data were acquired with different devices, post-processing was necessary to temporally register the simultaneous recordings with the same sampling rate and the same number of samples. To do so, randomly distributed triggers were sent to both kind of MEG sensors (SQUID or OPM) and SEEG. Data were then processed by an in-house Matlab routine to match the sequence of triggers and to resample the data (from SEEG to MEG time frame). The resampling procedure did not result in differences in delays larger than 1 sample. The end results for each session (SQUID-SEEG and OPM-SEEG) were two files that contained the same number of samples with synchronized SEEG and MEG data. Filtering: all data were band-pass filtered at 2-70Hz; a notch filter at 50Hz was added. SNR computation: to investigate the amplitude of events, the SNR was computed as the following ratio: the max amplitude of the event divided by the standard deviation of a baseline (2s of signal before the event for the single event presented in Fig. 2 and 500ms for the averages in Fig. 3 and Fig. 4).

**Fig. 2:**
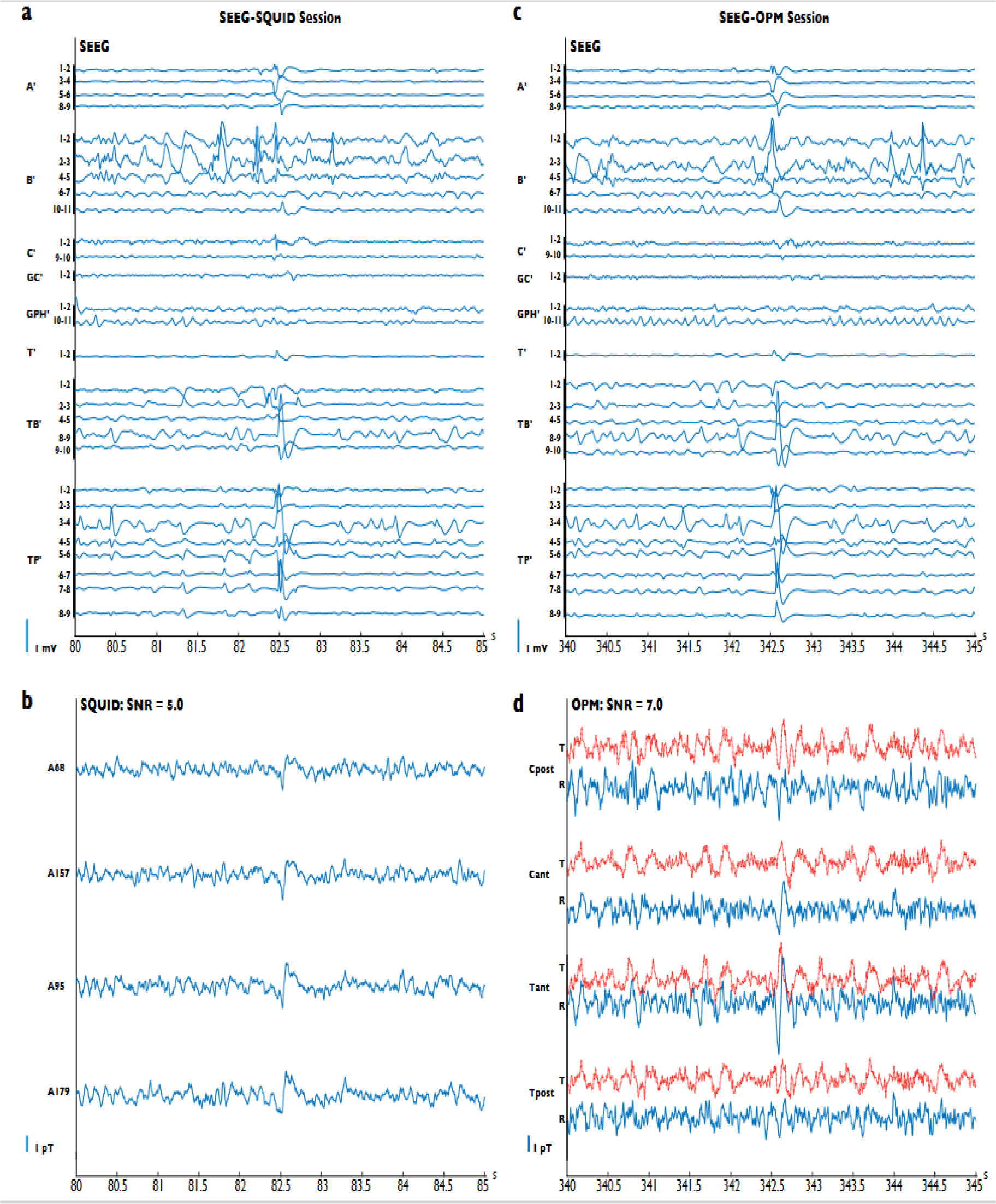
Individual spikes results. The left side of the figure presents the signal collected during the SQUID-MEG/SEEG simultaneous session: **a**. Bipolar SEEG data. The name of the SEEG electrodes and the recording contact label are shown. The labels range from 1 (deeper location) to 11 (most surface location). The spike clearly involves both deep and more superficial structures (amygdala, anterior and posterior hippocampus, third anterior temporal gyrus and temporal pole). **b**. Simultaneous SQUID-MEG data collected on the four sensors closest to the HeOPM channels: an interictal epileptic spike appears around 82.5s, with peak-to-peak amplitude of 1.1pT. The right side of the figure presents the signal collected during the ^4^He-OPM-MEG/SEEG simultaneous session. **c**. Bipolar SEEG data. Note the similarity between two intracerebral spikes disclosing the same anatomical location and time course. **d**. Simultaneous ^4^He-OPM-MEG data collected on four sensors: t=tangential magnetic field (red lines) and r=radial magnetic field (blue lines). A spike appears at 342.5s with a peak-to-peak amplitude of 2,5 pT. The vertical scale is identical to that of the SQUID data on the left.

**Fig. 3:**
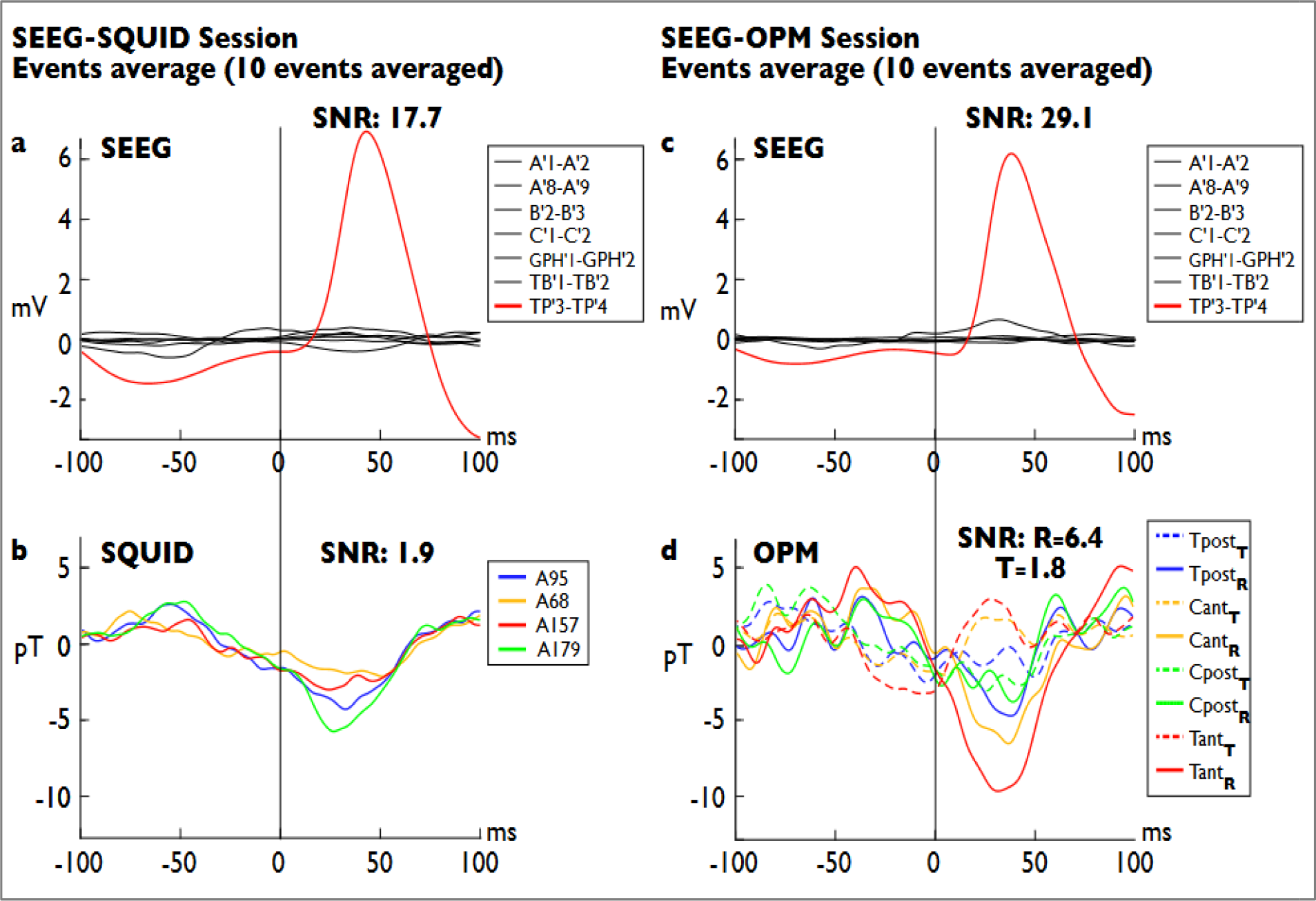
Averaged Type I spikes (n=10). The left side of the figure presents the averaged signal collected during the SQUID-MEG/SEEG simultaneous session, the right side presents the averaged signal collected during the ^4^He-OPM-MEG/SEEG simultaneous session. The name of the SEEG electrodes and recording contacts are shown. Contacts range from 1 (deeper location) to 9 (most surface location). **a**, Bipolar averaged SEEG data for the SQUID session. The averaged spike involves only the bipolar TP’3-TP’4 recording (Temporal pole). **b**, Simultaneous SQUID-MEG averaged data on the four sensors closest to the 4HeOPM sensors: a small interictal spike appears around 25ms with a peak-to-peak amplitude of 8 pT. **c**, Bipolar averaged SEEG data for the OPM session. Note the similarity between the intracerebral spikes of the two sessions, disclosing the same anatomical location and time course. **d**, ^4^He-OPM-MEG averaged data collected on four sensors (T_pos_, C_ant_, C_post_, T_ant_). The subscript indicates the orientation: T tangentia magnetic field (dashed lines), R radial magnetic field (solid lines). A spike appears at 35ms with a maximum peak-to-peak amplitude of 15 pT. The vertical scale is identical to that of the SQUID data on the left.

**Fig 4:**
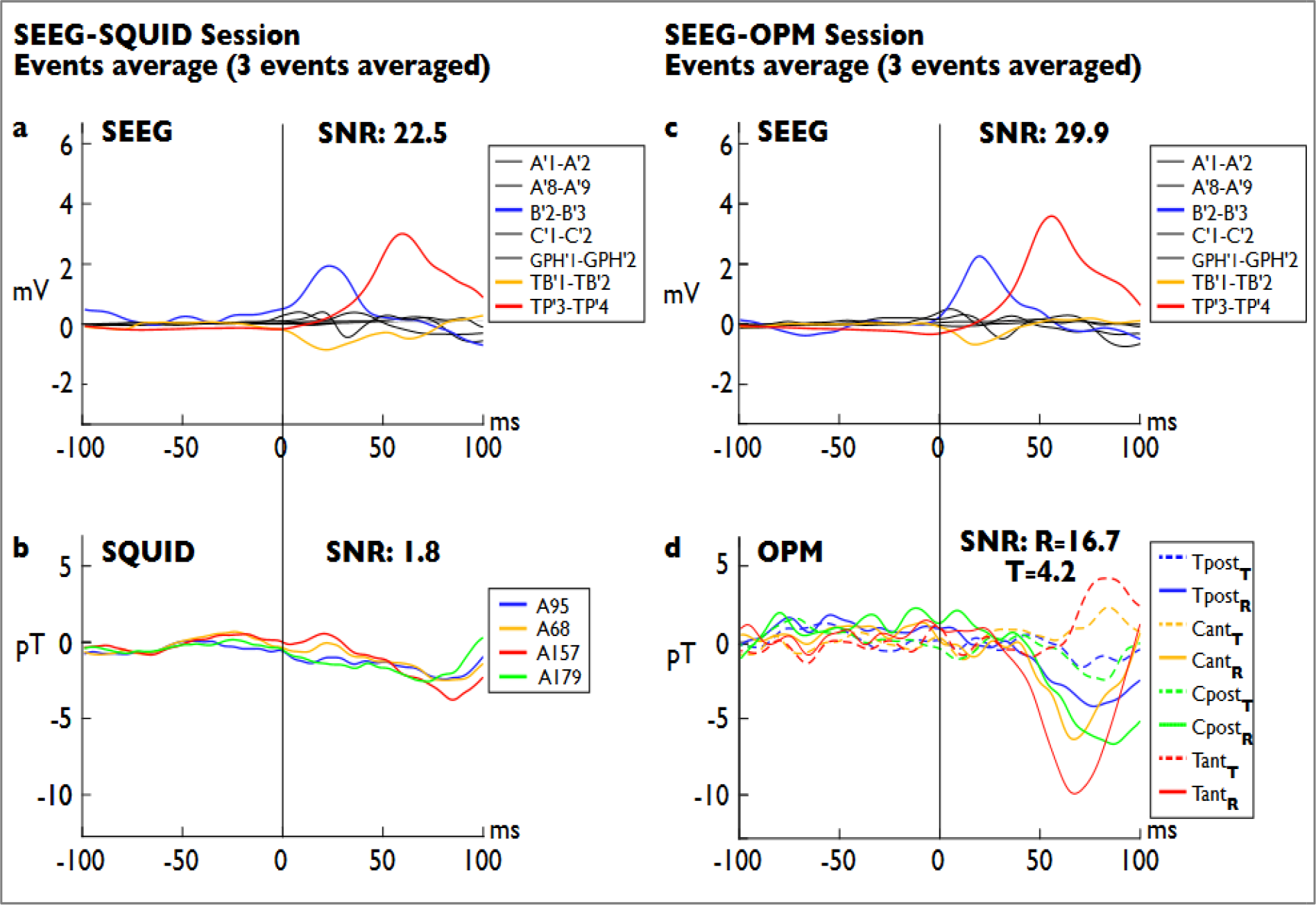
Averaged Type II spikes (n=3). Similar figure arrangement to that of Fig. 3. **a**, Bipolar averaged SEEG data for the SQUID session. The spikes involve a larger network encompassing TP’, TB’ and B’ electrodes (medial and lateral temporal pole, anterior hippocampus, third anterior temporal gyrus). **b**, Simultaneous SQUID-MEG averaged data on the four sensors closest to the 4He-OPM sensors: a very faint spike appears around 75ms with a peak-to-peak amplitude of 4 pT. **c**, Bipolar averaged SEEG data for the OPM session. Note the similarity between the intracerebra spikes of the two sessions, disclosing the same anatomical location and time course. **d**, 4He-OPM-MEG averaged data collected on four sensors (T_pos_, C_ant_, C_post_, T_ant_). The subscript indicates the orientation: T tangential magnetic field (dashed lines), R radial magnetic field (solid lines). A spike appears at 75s with a peak-to-peak amplitude of 9.3 pT maximum. The vertical scale is identical to that of the SQUID data on the left.

Automatic extraction of events of interest: We had to compare data recorded in the two separate sessions. To do so, we used a procedure to identify identical events across recordings. As a first step, an expert neurologist (FB) manually selected four kinds of events of interest from the SEEG recording of the SQUID session. The selection was done using a montage of one bipolar SEEG derivation per explored brain areas. These “reference” events were chosen to be representative of IEDs involving lateral temporal structures (Spike I) and IEDs involving both medial and lateral temporal structures (Spike II). We used the Matlab routine « findsignal » (Matlab 2020b) to find similar intracerebral events for both the SQUID session and the OPM session. Three to ten events were kept for averaging. From this set of events, we chose a representative occurrence of Spike II to illustrate a single event (See Fig. 1).

## Results

As a main result, the analysis of the simultaneous recordings globally shows that the activities recorded by the intracerebral electrodes placed in the left temporal pole are detected on the scalp surface by both systems, conventional SQUID-MEG and OPM-MEG (Fig. 2, 3, 4). Notably ^4^He-OPM-MEG recordings showed an SNR better than that of SQUID-MEG.

Single interictal events (epileptic spikes), are visible on both OPM and SQUID recordings. Fig. 2 presents single epileptic spikes with the corresponding SEEG traces. Using band-pass filtering (2 Hz to 70 Hz) and a notch filter (50 Hz) with no further processing, the ^4^He-OPM-MEG can clearly record, with a high signal-to-noise ratio (SNR), the IEDs identified with the SEEG electrodes. A single epileptic spike arising from the left temporal pole and the adjacent anterior third temporal gyrus (Fig. 2c shows the intracerebral traces), is distinctly identified on the scalp by the ^4^He-OPM-MEG (SNR=7, peak-to-peak amplitude=5pT) (Fig. 2d). In comparison, the four closest SQUID-MEG sensors, using advanced denoising based on reference sensors, detect an equivalent intracerebral spike (visible on simultaneous SEEG in Fig. 2a) but with a lower SNR of 5 (Fig. 2b). This can be explained by the reduced distance from the neuronal sources of ^4^He-OPM-MEG, yielding increased signal power compared to SQUID-MEG[Labyt et al., 2019]. Furthermore, it is interesting to note that different time courses and polarities are clearly identifiable, depending on the location of the channel and the orientation of the magnetic field (Fig. 2d). The ^4^He-OPM sensors are natively sensitive to two orthogonal orientations of the magnetic field, so that two different signals corresponding to the tangential and radial components (red lines and blue lines, respectively, in Fig. 2d) are output for each sensor.

These results observed on single spikes are even more evident when comparing the average of ^4^He-OPM-MEG and SQUID-MEG signals (Fig. 3). In order to compare a sufficient number of equivalent epileptic events recorded by OPMs and SQUID, we manually identified, using a selective SEEG electrodes montage, four different types of epileptic spikes recorded in the two separate simultaneous sessions (for details see Methods session and Supplementary Fig. 1 and 2). These IEDs have been subsequently automatically extracted from the entire time series and averaged by type. Two types of IEDs are illustrated, an average spike arising from the left temporal pole only (Fig. 3a and c, red line) and an average spike arising from the temporal pole, the third anterior temporal gyrus (minimally), and the anterior hippocampus (Fig. 4a and c, red, orange, and blue line, respectively). In both cases, the simultaneous SQUID-MEG averaged data reveal a small deflection (Fig. 3b and 4b, respectively -6pT and 3.7pT max value) in correspondence with the averaged intracerebral spikes (Fig. 3a and 4a). In contrast, the ^4^He-OPM-MEG averaged data disclose clear averaged spikes (Fig. 3d and 4d) occurring along with the two averaged intracerebral spikes (Fig. 3c and 4c). Both ^4^He-OPM-MEG spikes have a higher amplitude (spike I=-9.5 pT; spike II=-10pT) and a higher SNR (spike I: SNRr=6.4; spike II: SNRr=16.7) than those detected in the SQUID-MEG data. Regarding averaged spike-II, it is interesting to note that the time course of the OPM signal appears to be correlated with the decay of the temporo-polar spike, as recorded by electrode TP’ 3-4, whereas the hippocampal activity recorded by electrode B’ 2-3 is not detected by MEG sensors, neither by SQUIDs nor by OPMs. On the other hand, variable time-courses, polarities, and amplitudes between radial and tangential measurement axes can be observed on the OPM signals, particularly on spike II (Fig. 4d). The radial components of the two anterior sensors show a negative deflection while for the corresponding tangential components, the deflection is positive and slightly delayed. This suggests that ^4^He-OPM-MEG provides more information than SQUID-MEG about the spatio-temporal organisation of IEDs across the cerebral cortex thanks to their tangential component.

## Discussion

In this study we report the results of two sets of simultaneous intracerebral recordings of IEDs, one with ^4^He-OPM-MEG and the other one with SQUID-MEG. Using SEEG as reference and ground-truth, we correlated the IEDs recorded by intracerebral electrodes with both He-OPM and SQUID sensors. We obtained the first direct validation of the ability of ^4^He-OPM sensors to record epileptic activities and we demonstrate that new ^4^He-OPM system has better performance to record IEDs than SQUID-MEG system, as evidenced by its higher SNR. Notably, these recordings have been achieved in a regular clinical environment, without advanced noise correction. Thanks to their native 3D measurement of the magnetic field, OPM signals disclose variations in time-courses, polarities, and amplitudes between the radial and tangential components of the recorded activity.

Simultaneous acquisition of MEG and intracerebral EEG is a technical feat[Dalal et al., 2009; Dubarry et al., 2014; Santiuste et al., 2008] but can now be performed without major difficulties [Badier et al., 2017]. It has several key advantages in comparison with separate acquisition. It allows capturing the same activity at the surface and in depth, avoiding potential differences in brain state and medication. This is particularly important for IEDs that are spontaneous events which can widely vary in extent from one event to the other[Badier and Chauvel, 1995]. Simultaneity allows correlating signals across events[Dubarry et al., 2014] and in the current study, allowed finding similar events to be compared in SQUID and OPM sessions based on the SEEG topography. With this reliable comparison, we can establish that He-OPMs are at least as capable of recording epileptic activity as SQUIDs.

Currently, alkali-based OPM are mainly used for MEG recordings in healthy volunteers (for a review, see [Brookes et al., 2022]) and few studies in epileptic patients have been performed [Tierney et al., 2021; Vivekananda et al., 2020] [Feys et al., 2022], with one seizure recording recently reported [Feys et al., 2023]. Present results strengthen the perspective that OPMs are an accurate and valid alternative to SQUIDs.

Nonetheless, although very promising, clinical adoption of OPMs remains challenging due to some limitations of the current technology. Alkali OPMs have a limited bandwidth (1-100 Hz) and a small dynamic range (5 nT), requiring higher attenuation than a conventional shielded room for SQUID-MEG, demanding a complementary system of magnetic shielding coils to compensate for the remaining magnetic field and to reduce the cross-talk. Another challenge posed by the heat dissipation by sensors must be solved, possibly with a helmet design including insulation and/or cooling. Despite these constraints and thanks to technological advances to overcome them, alkali-based OPMs have given numerous proofs of their good sensitivity to biomagnetic measurements[Brookes et al., 2022; Labyt et al., 2019; Tierney et al., 2019]. As evidence of the rapid evolution of the technology, 90-channel OPM systems offering triaxial magnetic field detection have been shown to improve cortical coverage successfully [Boto et al., 2022; Rea et al., 2022]. The additional information offered by a vectorial measurement of the magnetic field, also reported in the present study, will be of interest to better characterize the spatio-temporal dynamics of the epileptic activity and to interpret clinical data [Iivanainen et al., 2017; Zahran et al., 2022]. In our study, the delay of the tangential component (see averaged spike-II) is potentially informative on the propagation of the interictal activity, as shown by comparing radial SQUID-MEG and EEG[Merlet et al., 1997].

Helium-based OPMs could overcome some of the limitations of alkali OPM, as they can be placed nearest the scalp without any discomfort for the patient,, since they do not require thermal insulation.. ^4^He-OPM are currently the only sensors to be self-compensated on their three measurement axes in a closed loop operating mode, ensuring a highly reliable measurement of the brain magnetic field, without cross-axis projection error[Fourcault et al., 2021], and bringing better stability of the sensors scale factor along time. This also allows for an increased dynamic range (up to 250 nT), larger than the one corresponding to the helium resonance linewidth, which eliminates the constraint of a strict field nulling system: in this study, ^4^He-OPMs have been used in a standard two-layer MSR in the hospital environment, known to be particularly magnetically “noisy”. On the other hand, in their current release,^4^He-OPMs have a worse sensitivity (40 fT/rtHz)[Fourcault et al., 2021] compared to alkali ones (20 fT/rtHz in tri axis mode)[Boto et al., 2022]. However, in a recent study in a large group of healthy subjects, the ^4^He-OPMs showed very similar results to the classical SQUID-MEG system thanks to their shorter distance to the brain[Gutteling et al., 2023]. In our study, we observed that ^4^He-OPM can record IEDs with a better SNR compared to SQUIDs. This result altogether with the recent data from 18 volunteers shows that ^4^He-OPMs are able to deliver high quality brain recordings[Gutteling et al., 2023].

Some limitation of this work comes from the limited number of OPM sensors. Nevertheless, the full coverage of the SQUID system allowed selecting of the co-localized SQUID sensors and ensuring a reasonable comparison. We found that the amplitude of the OPM signals was higher than that of SQUID signals, as expected from both simulations and real data[Boto et al., 2016; Boto et al., 2017; Iivanainen et al., 2017]. Still, the MEG IEDs, measured with 4He-OPMs and SQUIDs, arose from neocortical structures. We could not show activity from deep structures - such as the hippocampus - on either type of sensor. Sensitivity to deep sources remains a challenge. It will be necessary to use a larger number of sensors to verify if source separation techniques such as independent component analysis will allow to extract activity from deep sources as previously demonstrated[Pizzo et al., 2019].

With all its advantages, OPM technology could extend the use of MEG to many clinical and research applications. Currently, only very few clinical centres have a MEG facility. By offering more affordable cost, higher signal sensitivity and bandwidth, and portability without additional equipment, the OPM technology paves the way for the democratization of this unique non-invasive method of high-resolution brain exploration, potentially powerfully impacting both clinical practice and neuroscience.

## Data Availability

All data produced in the present study are available upon reasonable request to the authors

## Notes

### Competing Interest Statement

EL, APL, ML, holds founding equity in Mag4Health, a CEA-LETI spin off company whose aim is to commercialize Helium OPM based MEG systems. The remaining authors have no conflicts of interest.

### Funding Statement

This study was funded by AP-HM and CEA-LETI

### Author Declarations

The ethical approval was obtained at the AssistancePublique - Hopitaux de Marseille (AP-HM) under IDRCB 2020-A01830-39 and the patient gave informed written consent

